# Pacemaker implantation after cardiac surgery: a contemporary, nationwide perspective

**DOI:** 10.1101/2024.04.01.24305175

**Authors:** Amar Taha, Alice David, Sigurdur Ragnarsson, Piotr Szamlewski, Shabbar Jamaly, J. Gustav Smith, Susanne J. Nielsen, Anders Jeppsson, Andreas Martinsson

**Author notes:** **CORRESPONDING AUTHOR** Dr. Amar Taha, MD, Department of Molecular and Clinical Medicine, Institute of Medicine, Sahlgrenska Academy, University of Gothenburg, BOX 400, 405 30 Gothenburg, Sweden.

## Abstract

**Background:** Cardiac surgery entails an increased risk for brady-arrhythmias. Currently known rates for permanent pacemaker (PPM) implantation after cardiac surgery are derived from non-contemporary studies. This study aimed to explore the incidence and indications for PPM implantation at 30 days and one year after different cardiac surgical procedures in a modern cohort.

**Methods:** All first-time coronary artery bypass grafting (CABG) and/or valvular surgery patients in Sweden 2006 - 2020 without previous PPM or implantable cardioverter-defibrillator (ICD) (n= 76,447) were included in this nationwide population-based study using data from four national registries. Patients undergoing heart transplantation and those who subsequently received an ICD were excluded.

**Results:** A PPM was implanted in 8.2% (n= 6,271) of the patients, 35% (n= 2,196) within the first 30 days and 46.3% (n= 2,647) at one year following surgery. The main indication of PPM implantation was atrioventricular block. Tricuspid valve surgery exhibited the highest cumulative incidence for PPM both at 30 days (6.8% (95% confidence interval 4.3 – 10.0)) and one year (8.8% (95% CI 6.0 – 12.0)) surpassing mitral valve surgery (30-day 5.3% (95% CI 4.7-6.0); one-year 6.5% (95% CI 5.8-7.3)), aortic valve surgery (30-day 4.8% (95% CI 4.5-5.1); one-year 6.0% (95% CI 5.6-6.3)) and CABG (30-day 0.74% (95% CI 0.66-0.83); one-year 1.3% (95% CI 1.2-1.35)). The incidence following combined operations (multiple valves and/or coronary surgery) was 6.5% (95% CI 6.0 – 6.9) and 8.1% (95% CI 7.7 – 8.6) at 30 days and one year respectively. Concomitant ablation surgery increased the risk even further (adjusted Hazard Ratio 9.20 (95% CI 7.96-10.64), p<0.001).

**Conclusions:** The need for PPM after cardiac surgery is common, primarily due to atrioventricular block. Tricuspid valve surgery is associated with the highest risk for PPM implantation amongst isolated procedures. Combined procedures and concomitant surgical ablation further increase that risk.

**CLINICAL PERSPECTIVE:** *What is new?:*

- Permanent pacemaker after cardiac surgery is not uncommon with atrioventricular blocking being the main indication.
- Tricuspid valve surgery had the highest cumulative incidence at 30 days and one year, exceeding other isolated procedures.
- Combined cardiac surgical procedures and concomitant ablation surgery increased the risk even further.

*What are the clinical implications?:*

- This information is valuable while informing individual patients awaiting cardiac surgery about potential post-operative complications.
- Recognizing patients at higher risk enables targeted postoperative care, including closer monitoring for signs of conduction disturbances.
- Studies investigating and identifying predictors of post-cardiac surgery bradyarrhythmias and subsequent need for permanent pacemakers are needed.

## INTRODUCTION

Cardiac surgery is associated with a risk of non-transient bradyarrhythmia, requiring the implantation of a permanent pacemaker (PPM). (1) Due to the proximity of the heart valves to the atrioventricular node and the bundle of His, trauma during valvular surgery may lead to impaired electrical conduction between atrium and ventricle, atrioventricular block. (2) In addition, injury to the sinoatrial node may occur during lateral atriotomy or transseptal approaches targeting the mitral valve, resulting in sinus node dysfunction. (1) Alternatively, an underlying pathology could be unmasked by an inflammatory response during and after surgery, and/or extensive monitoring present after cardiac surgery leading to the diagnosis of a previously unknown conduction disorder.

Current estimates of PPM implantation following cardiac surgery are based on studies with data from the end of the 1990s or early 2000s, with considerable variations in the reported rates (from 0.8% to 9.7%). (3–5) Recent efforts addressing this issue have also yielded markedly variable results with data from either primarily earlier periods, single-centre experiences or with a focus on a single cardiac procedure. (1, 6–8) These discrepancies in reported rates of PPM after cardiac surgery are mainly due to heterogeneity in the selected population (e.g., valvular, or non-valvular heart surgery, PPM implantation in-hospital or post-discharge), different follow-up periods and study endpoints, and some studies including patients receiving an implantable cardioverter-defibrillator (ICD). (7) A large registry-based study from 2017 compared the risk of PPM implantation following valvular surgery versus coronary artery bypass grafting (CABG) over ten years. (9) However, patients who underwent mitral valve surgery or tricuspid valve annuloplasty were excluded from that study, and the underlying pathology leading to PPM implantation was not reported.

The aim of this study was to explore the PPM implantation rates and indications at 30 days, at one year, and beyond the first postoperative year following a wide range of cardiac surgical procedures in a large contemporary nationwide cohort. To further investigate the long-term risk for PPM implantation, the risk in cardiac surgery patients was compared to an age- and sex-matched cohort from the general population.

## METHODS

### Study design and data sources

The current study was an observational, nationwide study using prospectively collected data from four nationwide Swedish registries. The registries used in the current study were the Swedish Cardiac Surgery Registry, the National Patient Registry, the Swedish ICD & Pacemaker Registry, and the Swedish Total Population Registry.

The Swedish Cardiac Surgery Registry is part of the Swedish Web-system for Enhancement and Development of Evidence-based care in Heart disease Evaluated According to Recommended Therapies (SWEDEHEART) registry and has since 1992 contained detailed information on demographic and procedural characteristics for all cardiac operations in Sweden with high validity. (10–12) This registry was used to identify patients who underwent cardiac surgery and characterize the type of surgery performed.

Pre-procedural comorbidities were collected from the National Patient Registry which contains all hospital-based diagnoses registered according to the International Classification of Diseases, Tenth Revision (ICD-10) since 1987 for inpatient care and since 2001 in the outpatient setting. This registry has complete national coverage, and reporting to this registry is mandatory. (13) Information on the type of cardiac arrhythmia devices, date of implantation, and indication for implantation was obtained from the Swedish ICD & Pacemaker Registry, which has had an online platform since 2002 (http://www.pacemakerregistret.se). All centres in Sweden that implant cardiac implantable electronic devices report to the registry. The data in this registry is regularly monitored for internal validity. (14) Finally, the Swedish Total Population Registry (15) was used to extract the control population and mortality data. Individual patient data from the registries were linked using the unique personal identification number that all residents in Sweden possess.

The present manuscript was composed in accordance with the recommendations in the Strengthening the Reporting of Observational Studies in Epidemiology (STROBE) statement. (16)

### Study cohort

All adult patients in Sweden, with no history of previously implanted PPM or ICD, and who underwent first-time cardiac surgery between January 2006 and December 2020 were identified using the SWEDEHEART Registry and the Swedish ICD & Pacemaker Registry. The following groups of patients were excluded: those who underwent 1) heart transplantation, 2) any cardiac surgery procedure that did not include any stand-alone or combination of coronary artery bypass grafting (CABG), mitral valve surgery, aortic valve surgery or tricuspid valve surgery 3) subsequent ICD implantation. The final population comprised 76,447 patients undergoing CABG, mitral, aortic or tricuspid valve surgery. A flow chart depicting the included and excluded patients is presented in Figure 1.

**Figure 1.**
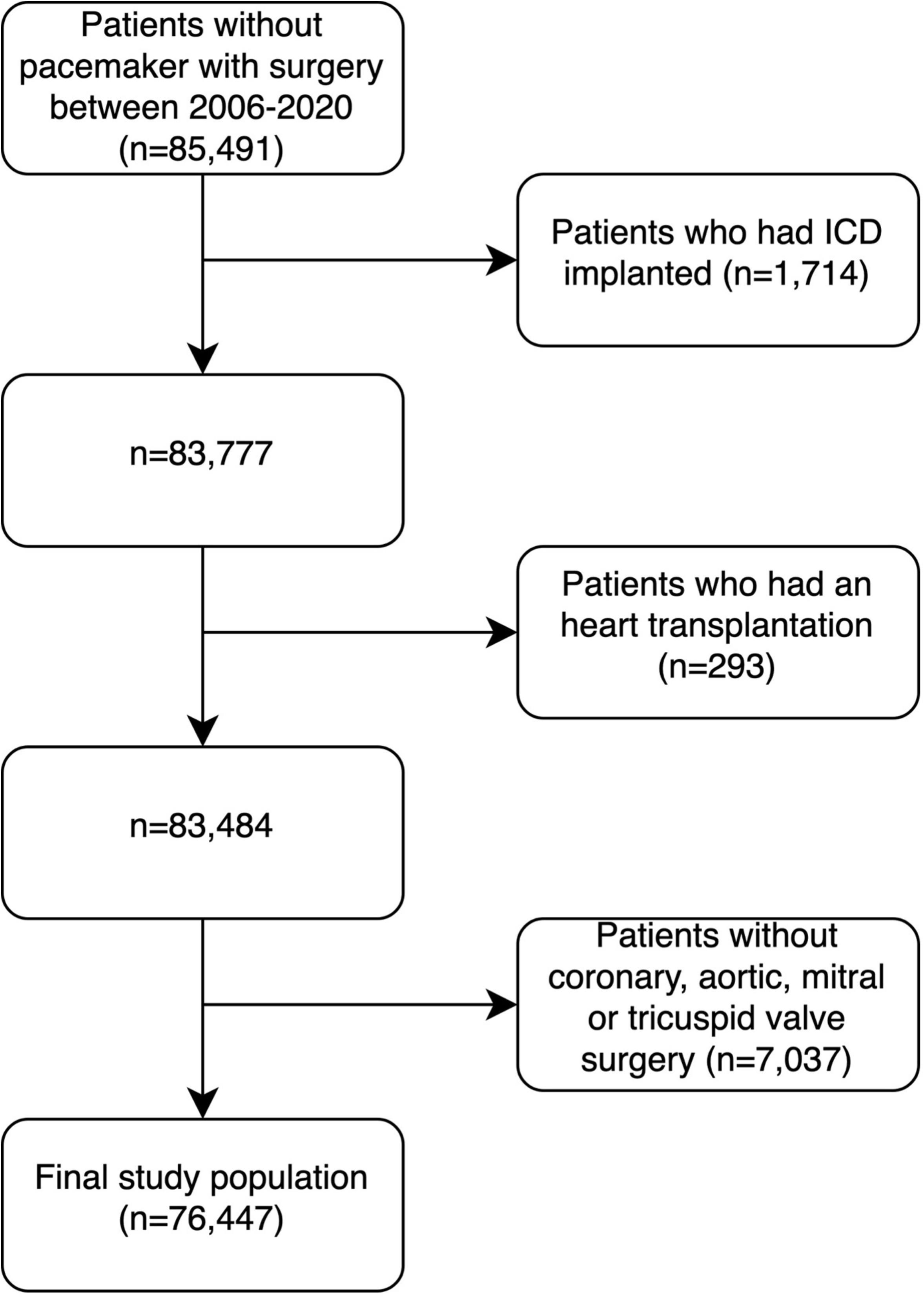
Flow chart of included and excluded patients. (ICD: Implantable cardioverter-defibrillator)

To evaluate the long-term risk of PPM after the first year, controls with no previous cardiac surgery were age- and sex-matched in a 1:1 fashion to the patients who underwent cardiac surgery. The date of inclusion of the controls was the date of the operation for the cardiac surgery patients. These controls were subsequently compared to patients who underwent cardiac surgery, both groups had to have survived the first postoperative year and had at least one year of follow-up. Patients with a previous pacemaker or pacemaker within one year after index surgery were excluded.

### Outcome measures

The primary study outcome was the cumulative incidence of pacemaker implantation at 30 days and one year. Secondary outcomes were pacemaker implantation at long-term follow-up and time from the cardiac surgical procedure to PPM implantation. In addition, the main indication for PPM implantation was explored. Using the control population, freedom from PPM implantation after the first postoperative year was compared between patients and control subjects. The one-year grace period was implemented to allow for the acute risk increase to regress to normal for patients who had cardiac surgery and subsequently evaluate the long-term risks after surgery compared to healthy controls.

### Statistical analysis

For baseline characteristics, continuous variables were either presented as means with standard deviation (SD) or medians with interquartile range (IQR) depending on their distribution. Categorical variables were presented as numbers and percentages. Point estimates for incidence rates were calculated by dividing the number of events by follow-up years. The corresponding 95% confidence intervals (95% CI) were calculated under the assumption of a Poisson distribution. To allow for competing risks, the cumulative incidence was used to assess incidence over time at 30 days, one year and 10 years. Competing risks regression was used to calculate subdistribution hazards for the risk of pacemaker implantation stratified per year between different types of procedures in the setting of competing risks and was estimated using Fine-Gray regression. Cox proportional hazard models were used to calculate adjusted hazard ratios (aHR) with 95% CI when comparing cardiac surgery patients with control subjects. Since the aim of the Cox regression analysis was to evaluate the long-term difference, the baseline was set at one year after surgery. All included patients and control subjects had to survive one year in this specific analysis. No patients had any missing data in that analysis. Logistic regression analysis adjusted for age- and sex was used to establish an association between the year of operation and incident PPM.

Patients with different combinations of cardiac procedures were evaluated both separately and in six main groups, patients could exclusively be assigned to one group: 1. Isolated CABG, 2. Isolated aortic valve surgery, 3. Isolated mitral valve surgery, 4. Isolated tricuspid valve surgery, 5. Combined valve surgery or combined valve surgery and CABG, 6. Concomitant arrhythmia surgery (independently of the primary procedure). Since arrhythmia surgery was almost exclusively combined with other procedures, we also performed a Cox regression analysis to evaluate the impact that arrhythmia surgery had on each of the other surgery groups. All tests were two-tailed and interpreted at the 0.05 significance level. All analyses were performed using R version 4.2.3 (R Foundation for Statistical Computing, Vienna, Austria).

### Ethical statement

The study complied with the declaration of Helsinki and was approved by the Swedish Ethical Review Authority (registration number 2021-00122, approved March 31, 2021), which waived the need for individual patient consent due to the observational register-based nature of the study.

## RESULTS

### Cohort description

A total number of 76,447 patients undergoing cardiac surgery were included. Median follow-up was 5.8 years (IQR 2.6-9.7). The mean age was 68 ((SD) 11) and 25.5% were females. Isolated CABG was performed in 41,946 (54.9%) patients, isolated mitral valve surgery in 4,695 (6.1%), 17,243 (22.6%) isolated aortic valve surgery, and 309 (0.4%) underwent isolated tricuspid valve surgery. Combined valve or valve and coronary surgery was performed in 9,867 (12.9%) of the patients, and 2,387 (3.1%) underwent concomitant surgical ablation (Table 1).

**Table 1.** Baseline characteristics.

|  | No<br>pacemaker<br>(N=73,544) | Pacemaker within<br>one year<br>(N=2,903) | Overall<br>(N=76,447) |
| --- | --- | --- | --- |
| **Sex** |  |  |  |
| Female | 18,603 (25.3) | 886 (30.5) | 19,489 (25.5) |
| Male | 5,4941 (74.7) | 2,017 (69.5) | 56,958 (74.5) |
| **Age (years)** |  |  |  |
| Mean (SD) | 68 (11) | 70 (11) | 68 (11) |
| **Body mass index (kg/m<sup>2</sup>)** |  |  |  |
| Underweight (<20) | 1,959 (2.7) | 122 (4.2) | 2,081 (2.7) |
| Normal (20–25) | 22,136 (30.1) | 1,015 (35.0) | 23,151 (30.3) |
| Overweight (25–30) | 32,382 (44.0) | 1,183 (40.8) | 33,565 (43.9) |
| Obese (>30) | 17,067 (23.2) | 583 (20.1) | 17,650 (23.1) |
| **LVEF** |  |  |  |
| >50% | 52,893 (71.9) | 1,959 (67.5) | 54,852 (71.8) |
| <50% | 20,651 (28.1) | 944 (32.5) | 21,595 (28.2) |
| **Heart failure** | 15,921 (21.6) | 1,021 (35.2) | 16,942 (22.2) |
| **Atrial fibrillation** | 26,808 (36.5) | 1,660 (57.2) | 28,468 (37.2) |
| **Prior stroke** | 6,495 (8.8) | 312 (10.7) | 6,807 (8.9) |
| **Prior myocardial infarction** | 26,344 (35.8) | 634 (21.8) | 26,978 (35.3) |
| **Chronic respiratory failure** | 8,210 (11.2) | 364 (12.5) | 8,574 (11.2) |
| **Renal failure** | 4,854 (6.6) | 308 (10.6) | 5,162 (6.8) |
| **Diabetes mellitus** | 18,119 (24.6) | 605 (20.8) | 18,724 (24.5) |
| **Hypertension** | 47,848 (65.1) | 1,836 (63.2) | 49,684 (65.0) |
| **Type of surgery** |  |  |  |
| Isolated CABG | 41,392 (56.3) | 554 (19.1) | 41,946 (54.9) |
| Isolated aortic valve surgery | 16,218 (22.1) | 1025 (35.3) | 17,243 (22.6) |
| Isolated mitral valve surgery | 4,391 (6.0) | 304 (10.5) | 4,695 (6.1) |
| Isolated tricuspid valve surgery | 282 (0.4) | 27 (0.9) | 309 (0.4) |
| Combined cardiac surgery | 9,145 (12.4) | 722 (24.9) | 9,867 (12.9) |
| Associated arrhythmia surgery | 2,116 (2.9) | 271 (9.3) | 2,387 (3.1) |

Number and proportion (%) or mean and standard deviation (SD).
CABG: coronary artery bypass grafting; LVEF: left ventricular ejection fraction

### Patient population

A PPM was implanted in 8.2% (6,271/76,447) of the patients during follow-up with 2,196/6,271 (35.0%) implantations occurring within the first 30 postoperative days and 2,903/6,271 (46.3%) within one year from the cardiac surgical procedure, thus the overall incidence of PPM implantation within one year was 3.8%. The median follow-up time from surgery to PPM implantation was 548 days (IQR 11-2207) for all patients receiving a pacemaker. The median time to PPM implantation was 10 days (IQR 7 – 34 days) for those who received a PPM during the first postoperative year. Baseline characteristics for patients with and without PPM at one year from cardiac surgery are presented in Table 1. Patients who received a PPM during the first postoperative year were generally older, more often female, had a lower left ventricular ejection fraction, and had other surgery than isolated CABG compared to patients who did not get a PPM within the first year. Heart failure, atrial fibrillation (AF), prior stroke, chronic respiratory disease, and renal failure were more frequent in patients who received a PPM whereas those without a PPM more often had myocardial infarction and diabetes mellitus.

### Incidence of PPM following cardiac surgery

Short-term cumulative incidence of PPM implantation, both at 30 days or one-year, was higher following isolated mitral valve (30-day 5.3% (95% CI 4.7-6.0); one-year 6.5% (95% CI 5.8-7.3)) and aortic valve surgery (30-day 4.8% (95% CI 4.5-5.1); one-year 6.0% (95% CI 5.6-6.3)) than after CABG (30-day 0.74% (95% CI 0.66-0.83); one-year 1.3% (95% CI 1.2-1.35)) (Figure 2). Isolated tricuspid valve surgery had the highest incidence of PPM implantation of the solitary procedures (30-day 6.8% (95% CI 4.3-10.0); one-year 8.8% (95% CI 6.0-12.0)). In patients who underwent a combined valve and/or coronary surgery the incidence of PPM was 6.5% (95% CI 6.0-6.9) and 8.1% (95% CI 7.7-8.6) at 30 days and one year respectively. The incidence of PPM increased over time in most groups of cardiac surgical procedures (Table 2). The highest incidence for PPM implantation was observed when concomitant ablation surgery was performed, and it increased the risk of PPM implantation in all combinations of procedures. A detailed description of the cumulative incidence of PPM implantation following different combinations of cardiac surgical procedures is presented in Supplemental Table S1. Estimates for PPM within one year calculated with cumulative risk regression adjusted for age and sex and with isolated CABG as reference are illustrated in Figure 2. Compared to isolated CABG, all other procedures had a significant and marked association with a higher risk for PPM implantation at one year, with arrhythmia surgery having the highest risk (adjusted HR 9.20 (95% CI 7.96-10.64), p<0.001).

**Figure 2.**
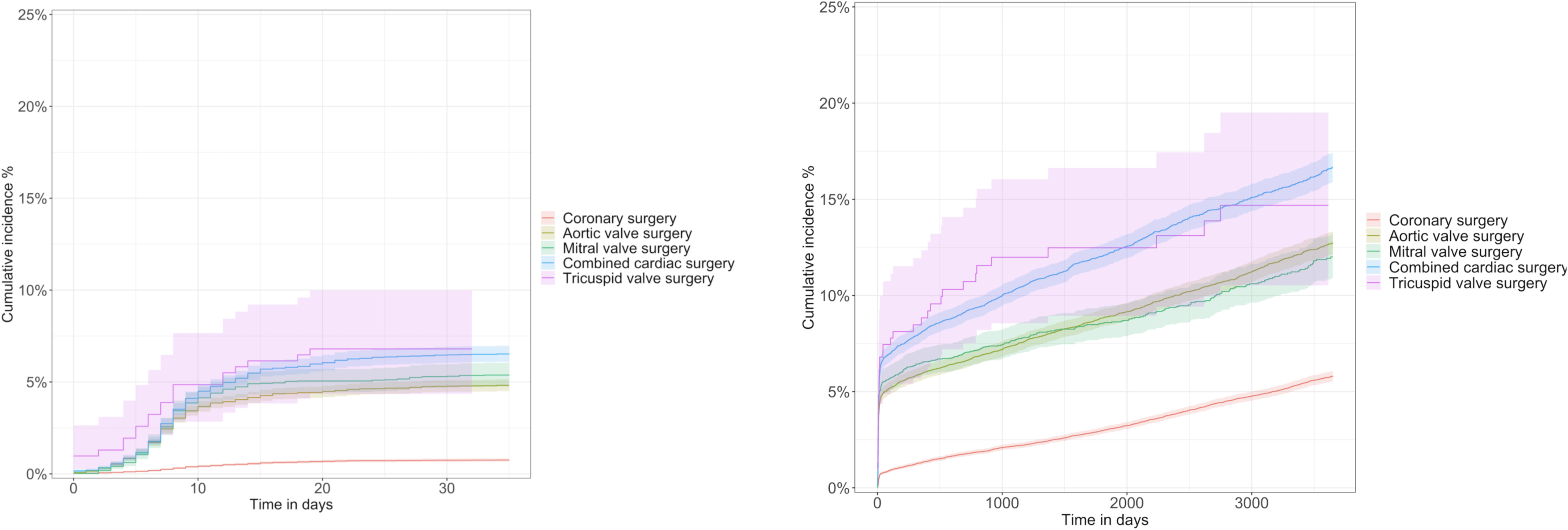
Cumulative incidence of permanent pacemaker implantation following cardiac surgery.

**Table 2.** The cumulative incidence (with 95% confidence interval) of permanent pacemaker (PPM) implantation at 30 days, one year and 10 years by type of cardiac surgery.

| Type of surgery | PPM at 30 days | PPM at one year | PPM at 10 years |
| --- | --- | --- | --- |
| Isolated coronary artery bypass grafting | 0.74%<br>(0.66–0.83) | 1.3%<br>(1.2–1.5) | 5.8%<br>(5.5–6.1) |
| Isolated aortic valve surgery | 4.8%<br>(4.5–5.1) | 6.0%<br>(5.6–6.3) | 12.5%<br>(12.0–13.0) |
| Isolated mitral valve surgery | 5.3%<br>(4.7–6.0) | 6.5%<br>(5.8–7.3) | 12.0%<br>(11.0–13.0) |
| Isolated tricuspid valve surgery | 6.8%<br>(4.3–10.0) | 8.8%<br>(6.0–12.0) | 15.0%<br>(11.0–20.0) |
| Combined valve and/or coronary surgery | 6.5%<br>(6.0–6.9) | 8.1%<br>(7.7–8.6) | 16.5%<br>(16.0–17.0) |

The 30-day cumulative incidence of PPM implantation for patients per calendar year is depicted in Supplemental Figure S1 and Supplemental Table S2. There was an increase in overall pacemaker implantation over the years (p<0.001). When the cumulative incidence was stratified based on the surgery group, the change was less apparent due to greater variability caused by a lower number of patients in each subgroup (Supplemental Figure S2).

In a cumulative risk regression analysis, stratified for each surgery group and adjusted for age and sex, operation in later years was associated with a higher incidence of PPM in patients with combined valve or valve and coronary surgery (HR 1.02 (95% CI 1.00-1.03 per year increase, p=0.016) and aortic valve surgery (HR 1.02 (95% CI 1.00-1.03 per year increase, p<0.001) but not for CABG surgery (HR 1.00 (95% CI 0.99-1.01) per year increase, p=0.78), concomitant surgical ablation (HR 1.01 (95% CI 0.98-1.03 per year increase, p=0.69) nor mitral valve surgery (HR 1.00 (95% CI 0.98-1.02) per year increase, p=0.81) or tricuspid valve surgery (HR 0.95 (95% CI 0.88-1.02)).

The risk for PPM implantation beyond the first postoperative year for patients who survived this period (n=66,581) was compared with age- and sex-matched controls (n=66,581) with no previous cardiac surgery. The hazard ratio for the risk of PPM implantation in patients with previous cardiac surgery was 3.75 (95% CI 3.50-4.01, Supplemental Table S3). When evaluating the addition of arrhythmia surgery to the different types of other surgery groups, an increased risk of future PPM implantation was noted for all surgery groups except for isolated tricuspid valve surgery which had similar point estimates but due to a smaller sample size, the confidence intervals were wide (Table 3). Isolated CABG was by far the most common surgery type but the largest burden of pacemaker implantation following surgery was from isolated aortic valve surgery (Figure 4), followed by combined valve and/or coronary surgery. Meanwhile concomitant arrhythmia surgery and tricuspid valve surgery, despite their higher incidence of pacemaker per procedure, were a less common cause of PPM due to their relative scarcity.

**Figure 3.**
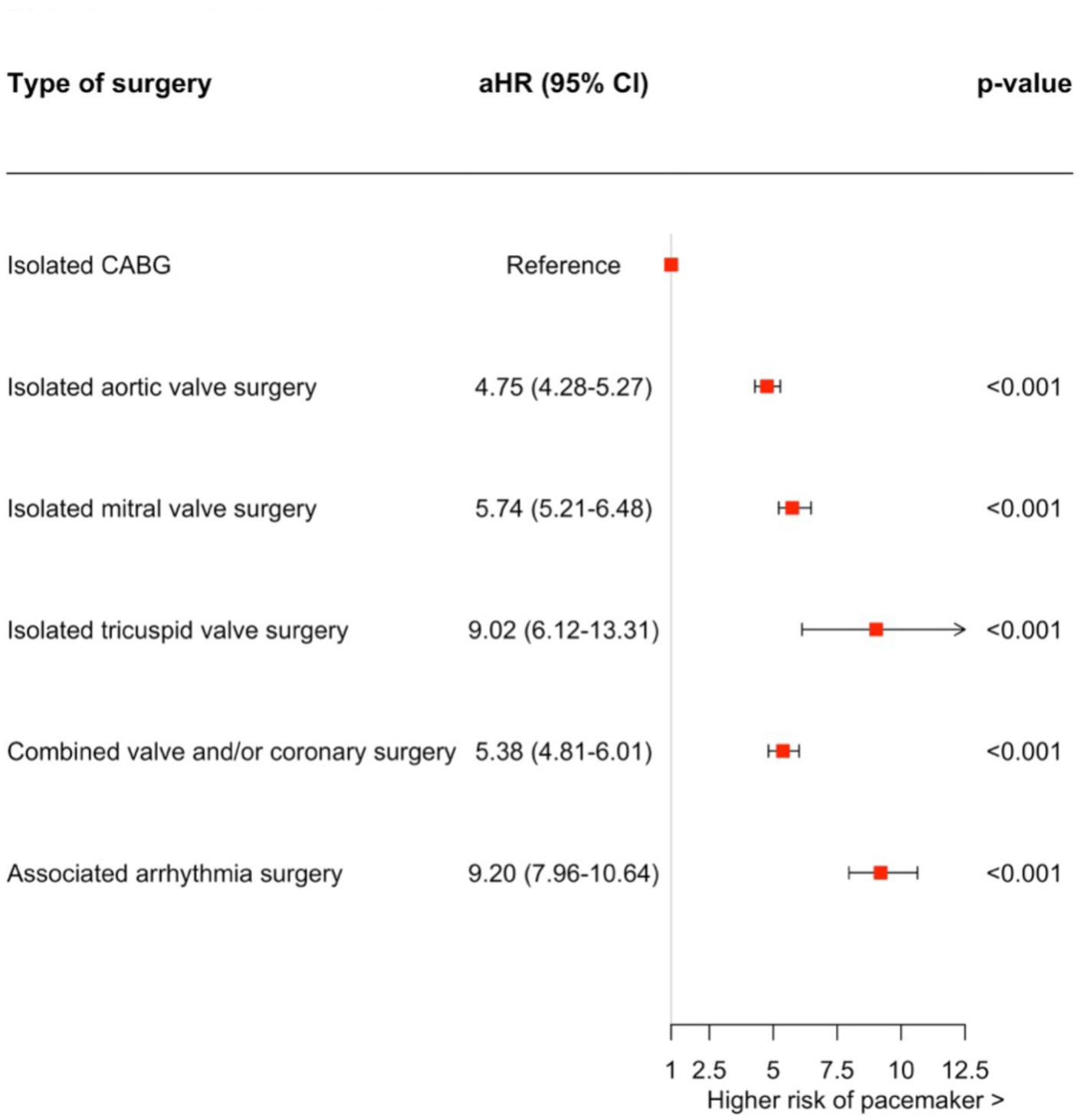
Forest plot illustrating the risk for permanent pacemaker, adjusted for age and sex, after different cardiac procedures with coronary artery bypass grafting as reference (aHR: adjusted hazard ratio; CI: confidence interval; CABG: Coronary artery bypass grafting)

**Figure 4.**
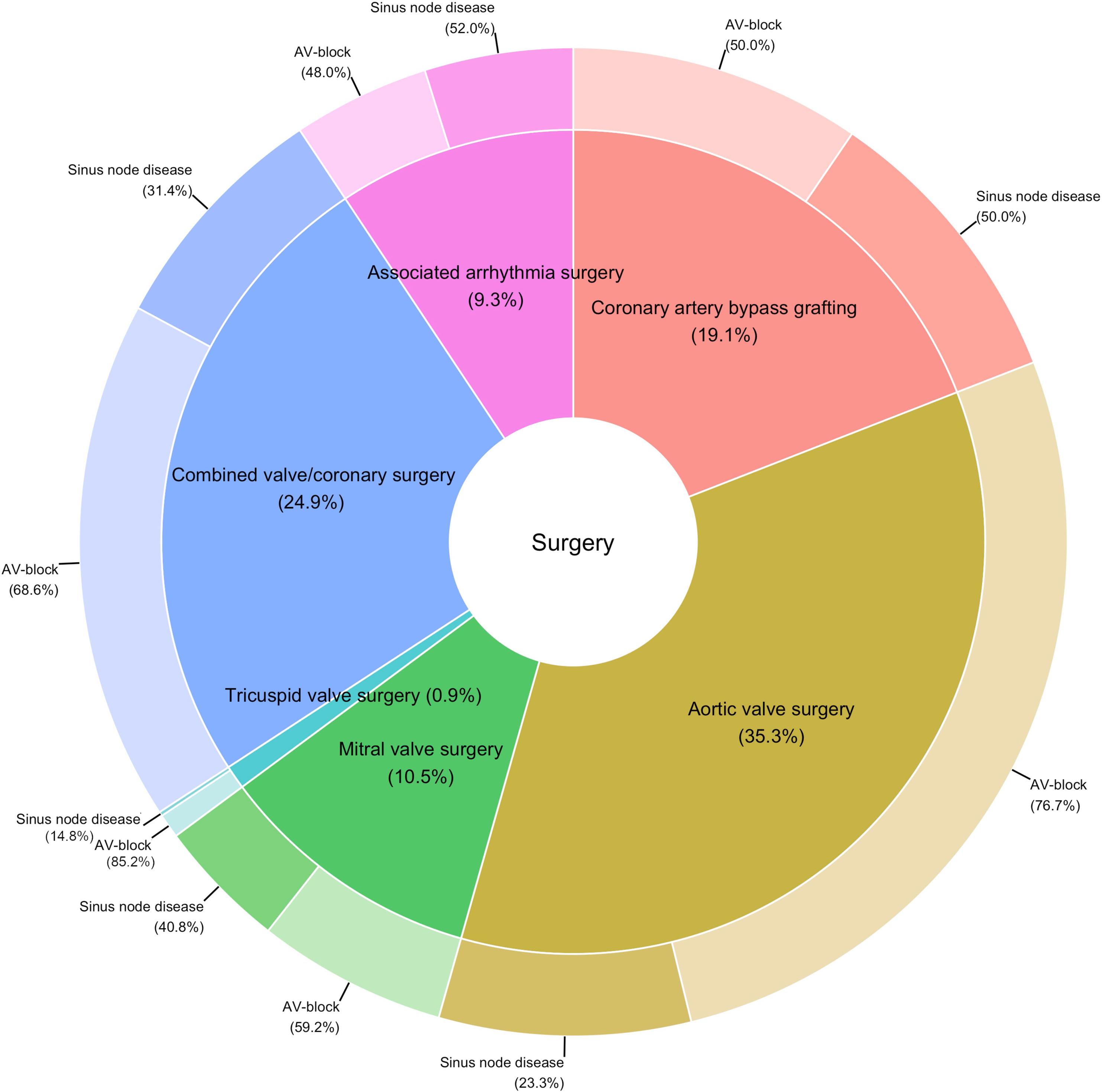
Distribution of pacemaker implantation during the first postoperative year by type of surgery and conduction system disease.

**Table 3.** Associations between addition of arrhythmia surgery and risk of pacemaker within one-year.

**Tabel Table 3. Associations between addition of arrhythmia surgery and risk of pacemaker** **within one-year**
| Type of surgery | aHR (95% CI) | p-value |
| --- | --- | --- |
| Isolated CABG | 2.44 (1.46-4.08) | <0.001 |
| Isolated aortic valve surgery | 1.87 (1.44-2.43) | <0.001 |
| Isolated mitral valve surgery | 1.54 (1.18-1.99) | 0.001 |
| Isolated tricuspid valve surgery | 2.00 (0.92-4.33) | 0.080 |
| Combined cardiac surgery | 2.55 (2.10-3.11) | <0.001 |
| Any surgery | 3.30 (2.91–3.74) | <0.001 |

Cox regression analysis adjusted for age and sex. CABG=Coronary artery bypass grafting.

### Indication for PPM implantation

Overall, atrioventricular block was the main indication in 65.1% of the patients in whom a PPM was implanted within the first postoperative year. Following isolated aortic valve, isolated mitral valve, and combined valve and coronary surgery, atrioventricular block was the predominant indication for PPM implantation. Sinus node dysfunction was the main indication for patients who underwent surgical AF ablation whereas following CABG, the distribution of these two PPM indications was similar (Table 4). Similar trends were found for those who had PPM implanted within 30 days.

**Table 4.** Indications for pacemaker implantation by type of cardiac surgery.

| Indication | Total<br>(N=2903) | Isolated<br>CABG<br>(N=554) | Isolated<br>aortic<br>valve<br>surgery<br>(N=1025) | Isolated<br>mitral<br>valve<br>surgery<br>(N=304) | Isolated<br>tricuspid<br>valve<br>surgery<br>(N=27) | Combine<br>d valve<br>and/or<br>coronary<br>surgery<br>(N=722) | Associated<br>arrhythmia<br>surgery<br>(N=271) |
| --- | --- | --- | --- | --- | --- | --- | --- |
| **30-days** |  |  |  |  |  |  |  |
| AVB | 1497<br>(68.2%) | 143<br>(46.1%) | 665<br>(80.8%) | 152<br>(61.0%) | 19<br>(90.5%) | 402<br>(69.9%) | 116<br>(53.2%) |
| SND | 699<br>(31.8%) | 167<br>(53.9%) | 158<br>(19.2%) | 97<br>(39.0%) | 2<br>(9.5%) | 173<br>(30.1%%) | 102<br>(46.8%) |
| **1 year** |  |  |  |  |  |  |  |
| AVB | 1891<br>(65.1%) | 277<br>(50.0%) | 786<br>(76.7%) | 180<br>(59.2%) | 23<br>(85.2%) | 495<br>(68.6%) | 130<br>(48.0%) |
| SND | 1012<br>(34.9%) | 277<br>(50.0%) | 239<br>(23.3%) | 124<br>(40.8%) | 4<br>(14.8%) | 227<br>(31.4%) | 141<br>(52.0%) |

AVB: Atrioventricular block; CABG: Coronary artery bypass grafting; SND: Sinus node disease.

## DISCUSSION

The main findings of this comprehensive, nationwide study are that the overall risk for PPM implantation within one year after cardiac surgery is 3.8%, mostly due to atrioventricular block, yet with large incidence variation depending on the type of index procedure performed. Amongst the isolated procedures, tricuspid valve surgery is associated with the highest risk of PPM, while combined valve and valve plus CABG, and concomitant surgical ablation are associated with the highest risk.

Previous research on the rate of PPM implantation post-cardiac surgery, largely based on data from the late 1990s and early 2000s, has shown considerable variability (3, 5). These studies were constrained by the type of surgical procedure included in the study population (4, 9), with limited follow-up periods (8), or including different types of cardiac implantable electronic devices in the studies (7). This present study stands out as the only contemporary study to comprehensively compare PPM implantation rates and indications across cardiac surgical procedures with complete follow-up. The findings from this study offer valuable insights for physicians and surgeons when the risk for PPM is discussed with individual patients awaiting cardiac surgery.

Amongst patients who underwent isolated procedures, tricuspid valve surgery was associated with the highest incidence of PPM implantation. One explanation for this finding is the proximity of the tricuspid valve to the atrioventricular node and bundle of HIS which could be affected by the surgical trauma leading to postoperative brady-arrhythmias. In addition, the indication for conducting isolated tricuspid valve surgery could be due to right-sided infectious endocarditis which could result in injury to the cardiac conduction system and lead to subsequent need for PPM.

Our study also highlights a 6.0% incidence of PPM implantation at one year following isolated aortic valve surgery. Aortic valve surgery was the most common surgery that required PPM due to a high caseload and relatively high frequency of PPM. Randomized controlled trials comparing transcatheter aortic valve replacement (TAVR), including The PARTNER 3 trial (17) and The EVOLUT trial (18), reported similar rates. Although in the PARTNER 3 trial, there was no significant difference in the need for new PPM implantation between patients who underwent surgical aortic valve replacement and those who underwent TAVR, other studies (18, 19) showed that TAVR was associated with a higher risk of PPM implantation. Given the increasing burden of valvular heart disease (20) and complications associated with PPM implantation, both in the short- and long-term (21, 22), the presence of preexisting conduction disturbances and the potential future need for PPM implantation in the individual patient merits consideration by the Heart Team when deciding on the type of intervention.

Another finding of note in our study was that patients who underwent concomitant surgical ablation had the highest risk of PPM implantation, with sinus node dysfunction being the main indication in this group which is in line with previous studies (8, 23). The association between concomitant surgical ablation and an increased risk of PPM implantation was notable in all types of surgery except for isolated tricuspid valve surgery. This might indicate that these patients are at a higher risk of pacemaker due to their underlying arrhythmic disorder, or that the procedure itself causes injury to the conduction system. The Society of Thoracic Surgeons 2017 guidelines for surgical treatment of AF recommend concomitant surgical ablation for AF at the time of mitral valve surgery (Class I recommendation, Level of evidence A). (24) Concomitant surgical AF ablation in patients undergoing valve surgery is also recommended by the European Society of Cardiology and the European Association for Cardio-Thoracic Surgery (Class IIa recommendation, Level of evidence A) (25). Studies comparing surgical and catheter ablation have however shown mixed results. The FAST (Atrial Fibrillation Catheter Ablation Versus Surgical Ablation Treatment) randomized 124 patients to either standalone surgical or catheter AF ablation. (26) Surgical ablation was superior to catheter ablation in achieving freedom from AF at 12 months. However, surgical ablation was associated with a higher complication rate, mainly driven by major bleedings and periprocedural pacemaker need. Later PPM implantation rates were not reported. Furthermore, 67% of patients included in that study had previously failed catheter ablation attempts which could possibly explain the higher success rate in the surgical ablation group. A more recent randomized trial (27) showed that catheter and surgical ablation were comparable in terms of efficacy with no patient in the catheter ablation group experiencing any adverse event in contrast to 20.8% adverse events occurring in the surgical ablation arm. Potential complications, including the risk for PPM implantation, should therefore be considered before surgical AF ablation.

The incidence of PPM implantation has risen over the study period, including following isolated aortic valve surgery, indicating that pacemaker implantations after cardiac surgery are unlikely to decrease over the coming years. This increase in the incidence of PPM is likely explained by the rapidly growing elderly population combined with an increase in the performance of cardiac surgery in elderly patients with higher frailty. (28) Finally, this risk for PPM implantation after the first postoperative year, compared to an age- and sex-matched cohort without a history of cardiac surgery, was more than three times higher. Despite that no adjustment for comorbid conditions was made, the intention of this analysis was merely to confirm that the risk for PPM implantation following cardiac surgery remains markedly elevated even beyond the first postoperative year. The current analyses cannot estimate if the increased HRs are caused by the residual risk associated with the surgery or, as is more likely, that it is caused by the prevalence of concomitant heart disease and comorbidity in the surgery group.

## LIMITATIONS AND STRENGTHS

Data on preexisting conduction abnormalities, such as bundle branch block or low-degree atrioventricular conduction disturbance were not available in the nationwide registers used here. Furthermore, information on the use of certain medications (e.g., beta-blockers or antiarrhythmic drugs), which could influence the risk for PPM implantation, was lacking. This study, however, outlines the real-world rates of PPM implantation after cardiac surgery across multiple centres in a large contemporary nationwide cohort using data collected from high-quality registries with complete coverage due to mandatory registration.

## CONCLUSIONS

The need for PPM after cardiac surgery is common and mostly due to atrioventricular block. Amongst isolated cardiac procedures, tricuspid valve surgery is associated with the highest risk for PPM, and both combined cardiac procedures and concomitant surgical ablation further increase that risk.

## Data Availability

The data used in this study will be provided upon reasonable request and following approval by the SWEDEHEART Registry, the Swedish National Board of Health and Welfare, and the Ethical Review Authority.

## ABBREVIATIONS

AVB: Atrioventricular block
CABG: Coronary artery bypass grafting
ICD: Implantable cardioverter defibrillator
PPM: Permanent pacemaker
SND: Sinus node dysfunction

## FUNDING

This work was funded by the Swedish state under the agreement between the Swedish government and the county councils concerning economic support of research and education of doctors (ALF agreement, ALFGBG-977905 to AM)

## DISCLOSURES

Anders Jeppsson discloses financial relationships with AstraZeneca, Werfen, and LFB Biotechnologies unrelated to the present study. Amar Taha discloses financial relationships with Medtronic and Abbott.

## Notes

### Competing Interest Statement

The authors have declared no competing interest.

### Clinical Trial

The study was not registered due to the retrospective and observational nature of the study.

### Author Declarations

The study was approved by the Swedish Ethical Review Authority (registration number 2021-00122, approved March 31, 2021).

